# Distinct Features of Predictive Profiles for Post-TAVR Functional Improvement and Mortality. Value of 6-minute Walk Test and Myocardial Work Analysis

**DOI:** 10.1101/2025.11.13.25340201

**Authors:** Jerzy Rychlewski, Monika Przewlocka-Kosmala, Thomas H. Marwick, Jonathan Sen, Krzysztof Aleksandrowicz, Grzegorz Golanski, Karolina Kubisz, Erwan Donal, Marcin Protasiewicz, Piotr Kubler, Wojciech Kosmala

## Abstract

Clinical improvement and survival benefit after transcatheter aortic valve replacement (TAVR) are difficult to predict. Despite the identification of multiple predictors of both endpoints, prognostic algorithms in patients undergoing TAVR are not well defined.

**Aim:** To explore factors determining the lack of significant improvement in exercise capacity after TAVR, and investigate whether the predictive profile for poor functional response to TAVR (PFR) could also predict mortality.

**Methods and results:** We enrolled 220 patients (78.0±6.4 yrs) with severe aortic stenosis who underwent TAVR. Pre- and post-procedural (3 months) data were collected, including clinical information, echocardiography with myocardial work assessment, and 6-minute walk test (6MWT). PFR was defined as an increase in 6MWT distance <40m.

Only age and atrial fibrillation were independent predictors of both mortality and PFR. Mortality was associated with lower global work index (HR 0.99; p<0.001) and shorter 6MWT distance (HR 0.99; p=0.008). PFR was associated with higher global wasted work (OR 1.003; p=0.026) and less impaired 6MWT relative to predicted values (OR 0.19; p<0.001 for 6MWT <80% predicted). Unsupervised machine learning-based analysis (Partition Around Medoids algorithm) identified clusters associated with mortality but not with PFR.

**Conclusions:** The predictive profiles for PFR and mortality after TAVR show only limited overlap, indicating that these two outcomes warrant distinct prognostic evaluations. The 6MWT may be useful in predicting the likelihood of both outcomes, with lower absolute values being associated with mortality, while higher percentage predicted values – with limited symptomatic benefit. Myocardial work analysis may further enhance prognostic assessment in this population.

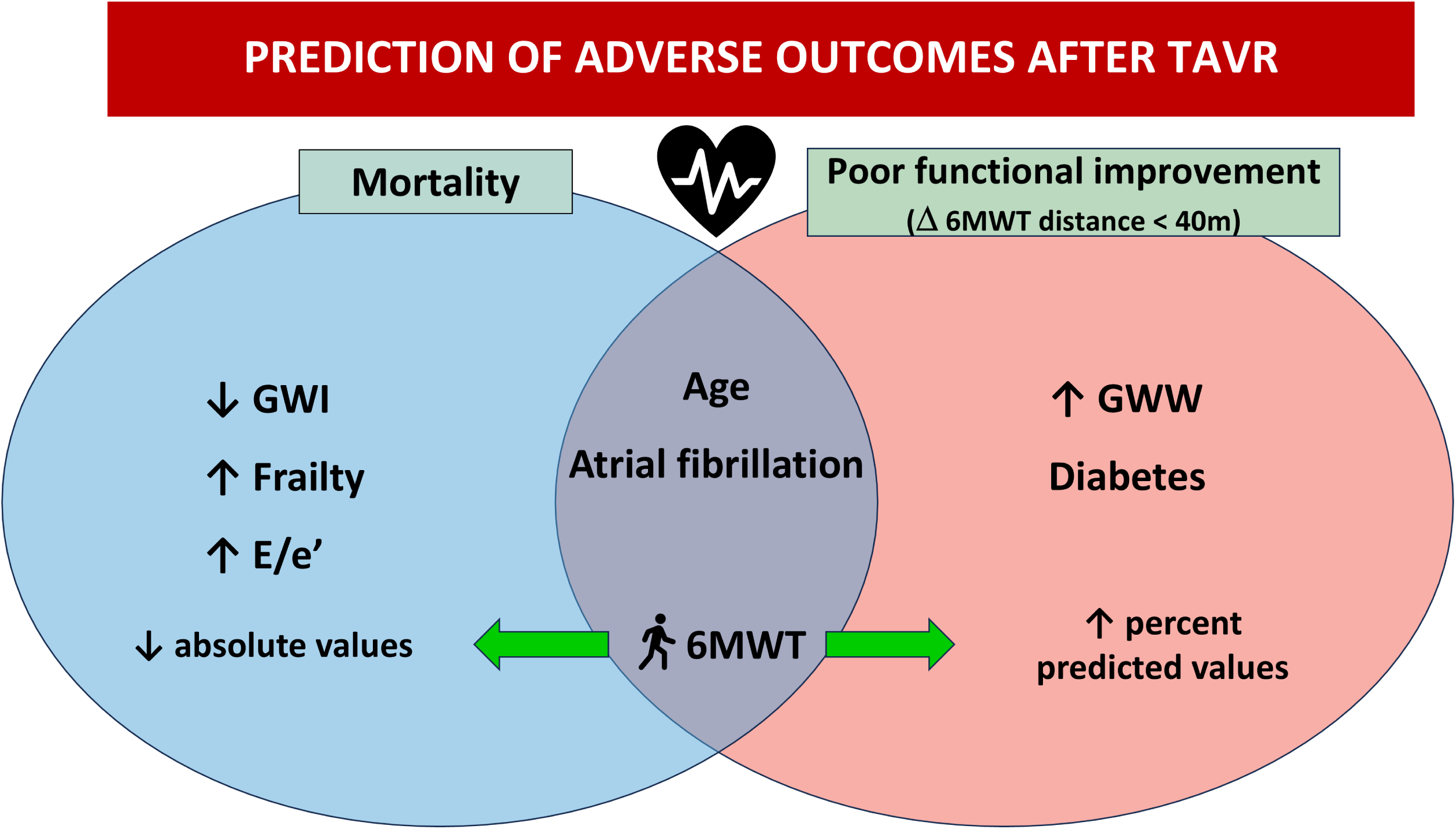

---

Transcatheter aortic valve replacement (TAVR) has become the treatment of choice for many patients with severe aortic stenosis (AS), with the number of procedures in the USA exceeding the volume of surgical aortic valve replacement and projected to reach 190 000 annually by 2029^1^. While TAVR offers clear benefits for patients at high surgical risk, it may prove futile in others. Many elderly and multimorbid individuals are referred for TAVR, among whom clinical improvement and survival benefit with this procedure may be uncertain. A significant proportion of patients undergoing TAVR experience no symptom relief after the intervention, and in some series, >25% of patients in high-risk subgroups die within 1 year^2–6^. Multiple factors may contribute to poor response, including frailty and comorbidities^4–6^, as well as irreversible myocardial fibrosis and cardiomyocyte damage resulting from longstanding AS^7, 8^.

Defining phenotypic features that characterize poor clinical responders to TAVR is important for both patient care and health economics. This diagnostic work-up should be based on commonly available resources to permit its widespread use. Key tools for gathering relevant information to address these issues include echocardiographic imaging, laboratory markers, and the 6-minute walk test (6MWT) that integrate cardiopulmonary and musculoskeletal function and correlate with peak oxygen consumption^9^. However, data supporting the prognostic utility of 6MWT after TAVR are scarce^10^, and guidelines for the management of valvular heart disease do not include this test in the diagnostic portfolio. Likewise, assessment of myocardial work from incorporating left ventricular (LV) afterload into global longitudinal strain (GLS) analysis could be an attractive method to track changes in cardiac mechanics after correction of AS and stratify clinical risk^11^. However, its diagnostic and prognostic role in this patient population has yet to be clearly established. Accordingly, we sought to investigate factors determining the lack of significant improvement in exercise capacity after TAVR despite favorable hemodynamic changes achieved with the intervention, with special focus on the utility of 6MWT and myocardial work assessment. In addition, we aimed to investigate whether the predictive profile for poor functional response to TAVR could also prove useful in predicting mortality, thus serving as a uniform prognostic tool in candidates for TAVR.

## Methods

### Patient selection

The study group of 220 patients satisfying the diagnostic criteria for symptomatic severe AS were prospectively selected from 249 candidates scheduled for TAVR between 2020 and 2022 in hospital clinics at a tertiary cardiology center. The diagnosis of severe aortic stenosis was ascertained by resting echocardiography (mean gradient ≥40 mmHg, peak velocity ≥4.0 m/s, valve area ≤1 cm^2^ [or ≤0.6 cm^2^/m^2^]), and in case of low-flow, low-gradient conditions - by low-dose dobutamine stress echocardiography, or calcium score in cardiac computed tomography as per recommendations^12^. Exclusion criteria encompassed insufficient image quality, severe aortic regurgitation and more than mild mitral valve disease on pre-TAVR echocardiography (**Figure 1**). The decision to undertake transcatheter aortic valve implantation was made after careful consideration of clinical, anatomical, and procedural factors by the multidisciplinary Heart Team. All participants were informed of the purpose of the study and provided written informed consent. Investigations were in accordance with the Declaration of Helsinki and were approved by the institutional ethics committee.

**Figure 1.**
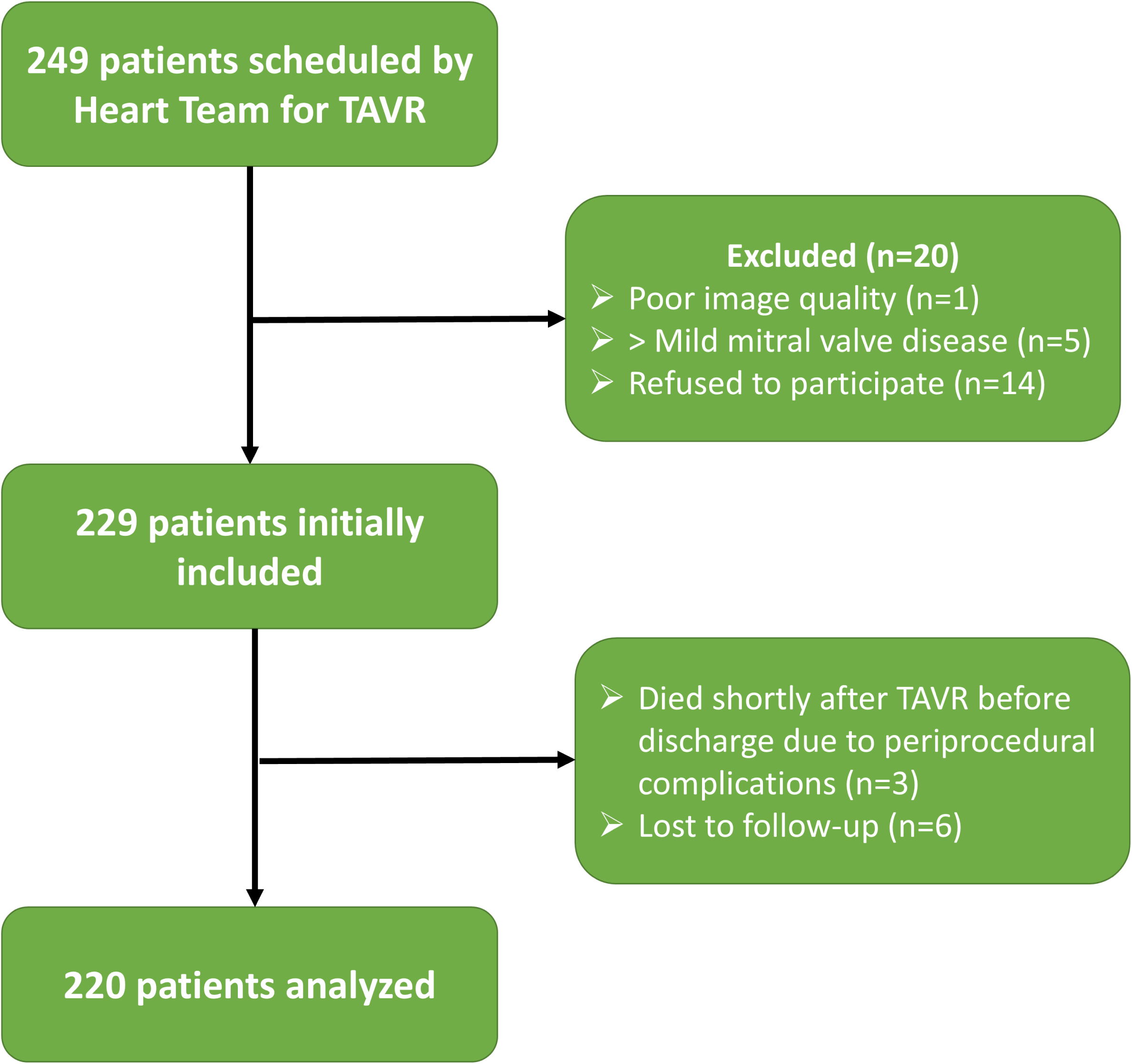
Patient flow diagram.

A group of 171 patients with AS scheduled for TAVR, meeting the same eligibility criteria as the studied population, served as the validation cohort. The clinical characteristics of this subset are presented in Supplementary Table 1. Given the limited follow-up duration in this subset (median of 12 months), the analysis focused solely on poor functional response to TAVR, without evaluating mortality.

### Study protocol

Study procedures including echocardiography, 6MWT, and blood sampling for laboratory analysis including NT-proBNP were performed twice: 2-4 weeks prior to TAVR and 3 months after TAVR. During evaluation, clinical data were collected, brachial blood pressure readings were taken, and resting electrocardiograms were recorded. Patient frailty was assessed using the Clinical Frailty Scale evaluating specific domains including comorbidity, function, and cognition^13^. The severity of a patient’s comorbidities was evaluated by the Charlson Index Score^14^.

### Study outcomes

To address the study aims, 2 separate outcome measures were assessed: poor functional response to TAVR and all-cause mortality. The threshold for significant improvement in 6MWT distance was set at 40 m based on available physiological data^15, 16^. The rate of mortality was verified by contacting patients or their proxies, and using the electronic population registration system.

### Echocardiography

Echocardiographic imaging was performed using standard equipment (Vivid E95, GE Medical Systems, Horten, Norway). The same imaging protocol was used at pre- and post-TAVR examinations and carried out by the same sonographer.

### Conventional and tissue Doppler imaging

Cardiac dimensions and wall thicknesses were measured according to established recommendations^17^. LV volumes with ejection fraction and LA volume were evaluated by the biplane Simpson method. The mitral inflow and mitral annular parameters were assessed in the apical four-chamber view. The E/e’ ratio was calculated by dividing the early diastolic mitral inflow velocity by the average early diastolic tissue velocity obtained from the septal and lateral parts of the mitral annulus.

Trans-aortic mean and maximum pressure gradients were measured using continuous wave Doppler. Aortic valve area was estimated with the continuity equation. Right ventricular function was assessed using tricuspid annular plane systolic excursion (TAPSE). Tricuspid regurgitant pressure gradient was used as an estimate of systolic pulmonary artery pressure.

### Speckle tracking imaging

A semiautomated two-dimensional speckle tracking technique (Echopac PC, General Electric Medical Systems) was used to measure longitudinal myocardial deformation in the three apical views, with a common temporal resolution of 60 to 90 frames per second. The peak negative value on the strain curve during LV systole was measured, and the average longitudinal deformation from the three apical views was reported as global longitudinal strain (GLS).

Myocardial work was assessed by integrating LV longitudinal strain data with noninvasively estimated LV pressure, derived from the sum of brachial systolic blood pressure (measured in the left lateral position during echocardiography) and the mean transaortic pressure gradient^18, 19^. Timings of mitral and aortic valve closure and opening were obtained from continuous-wave Doppler recordings and 2D images in the apical 3-chamber view. The following parameters were calculated: global myocardial work index (GWI), global constructive work (GCW), global work waste (GWW), and global work efficiency (GWE) as previously described^20^.

In patients with atrial fibrillation, LV longitudinal strain, myocardial work and ejection fraction were assessed using the index beat method, based on the extraction of 2 consecutive beats of similar duration, with an RR1/RR2 ratio ranging from 0.96 to 1.04^21^. Two-dimensional loops spanning a 15-second timeframe were recorded in each apical view to capture index beats with uniform RR intervals across the views, which was a prerequisite for the software’s approval to calculate myocardial work.

### 6-minute walk test

The 6-minute walk test was conducted by a team of physiotherapists according to a standardized protocol, using an internal corridor 25 meters long. Every participant was told to walk the longest possible distance at their own pace, within 6 minutes. During the test, patients could take a pause for a rest and restart as soon as they could^22^. The predicted distance of 6MWT was calculated from the standard Enright and Sherrill formula^23^.

### Statistical analysis

Data are presented as mean ± SD for normally distributed variables, as median (interquartile range) for skewed variables, and as counts and percentages for categorical variables. The association of demographic, clinical and echocardiographic features in patients with post-interventional change in 6MWT distance < and > 40 m were made using univariable logistic regression. Associations of survival were sought with univariable Cox regression.

#### Clustering Algorithms

A machine learning strategy was used because of the highly-dimensional nature of the data. Unsupervised learning methods based on clustering were applied to the 57 clinical variables (see Supplementary Materials for the full list of predictors) into meaningful clusters without using outcome data. The following clustering algorithms were tested: K-means, partition around medoids (PAM), agglomerative hierarchical clustering (complete-linkage), density-based spatial clustering of applications with noise (DBSCAN), and hierarchical density-based spatial clustering of applications with noise (HDBSCAN). The optimal number of clusters was initially estimated using two cluster validation metrics: silhouette width and Davies-Bouldin score. However, the final decision on the number of clusters was made by assessing the clinical interpretability of the clustering outputs. The clinical relevance of the clusters was prioritized, and the grouping that made the most sense based on domain knowledge was selected. Details of the machine learning methods with references are provided in the Supplementary Materials.

#### Post-hoc Interpretation of Clustering Analysis

To interpret the clustering results, variable importance was compared using the means of continuous variables and proportions of categorical variables. Model-Agnostic Explanation with SHapley Additive exPlanations (SHAP) were used to assess the importance of each variable in predicting cluster membership. SHAP values were calculated using Extreme Gradient Boosting (XGBoost). The top variables contributing to each cluster were identified and compared between clusters to understand which features were most important for defining the cluster structure.

#### Supervised Analysis for Phenotypic Differentiation

Following the unsupervised clustering, a supervised analysis was performed to investigate the relationships between clusters and clinical outcomes using logistic regression. Patient clusters were sought with machine learning, and differences in individual variables were sought using one-way ANOVA for continuous variables and chi square test for categorical variables.

Homogeneity of variances was assessed by implementing the Levene test. Variables with p values <0.2 in univariable associations with outcomes were tested in multivariable logistic and Cox regression models to define the best independent predictors of poor functional response to TAVR and mortality. Nested logistic regression models were built to evaluate the incremental predictive value of 6MWT (used as a binary parameter categorized based on a cut-off value of 80% predicted) and global wasted work for prediction of poor functional response to TAVR. The change in overall log-likelihood ratio chi-square was used to evaluate the improvement in predictive power after adding subsequent parameters. The Harrell c-statistic was used to assess model performance. Model R-squares were calculated to evaluate the proportion of variation in the outcome variable explained by the model. Skewed variables were subjected to a logarithmic transformation prior to their analysis. Post-TAVR changes in particular parameters were computed by subtracting the pre-TAVR value from the post-TAVR value and were expressed in the units of their measurements.

Calculations were carried out with standard statistical software (Statistica version 13.3, TIBCO Software Inc., Palo Alto, CA, USA). The clustering analyses and supervised analysis for phenotypic differentiation were performed in the R statistical platform (version 4.3.3, 2024-02-29) using the library packages: *dplyr, DesTools, tidyverse, lubridate, table1, flextable, randomForest, ggplot2, cluster, factoextra, clusterSim, dbscan, gtsummary, gt, shapper, SHAPforxgboost, xgboost, rms,* and *survival*.

## Results

### Patient characteristics

The clinical profile of 220 subjects enrolled in the study was characterized by advanced age typical of TAVR populations, a slight female predominance, a low to moderate burden of comorbidities except hypertension, relatively mild frailty, and significantly impaired exercise tolerance (**Table 1**).

**Table 1.** Demographic, clinical and echocardiographic characteristics of the studied population according to post-TAVR improvement in 6MWT.

| Parameter | All<br>n=220 | Group A<br>delta 6MWT>40 m<br>n=109 | Group B<br>delta 6MWT<40 m<br>n=111 | P (A-B) |
| --- | --- | --- | --- | --- |
| **Demographic and clinical parameters** |  |  |  |  |
| Age, years | 78.0±6.4 | 77.2±6.1 | 78.9±6.6 | **0.048** |
| Male sex, n (%) | 101 (46) | 55 (50) | 47 (42) | 0.227 |
| Atrial fibrillation, n (%) | 32 (15) | **7 (6)** | **25 (23)** | **<0.001** |
| Pacemaker after TAVR, n (%) | 18 (8) | 7 (6) | 11 (10) | 0.345 |
| Coronary artery disease | 116 (53) | 56 (51) | 60 (54) | 0.691 |
| Concomitant PCI, n (%) | 43 (20) | 19 (17) | 24 (22) | 0.433 |
| Hypertension, n (%) | 199 (90) | 97 (89) | 102 (92) | 0.464 |
| Diabetes, n (%) | 91 (41) | 38 (35) | 53 (48) | 0.052 |
| Lung disease, n (%) | 32 (15) | 18 (17) | 14 (13) | 0.411 |
| Systolic blood pressure, mmHg | 136±18 | 29.2±5.7 | 29.0±4.9 | 0.815 |
| Diastolic blood pressure, mmHg | 76±11 | 136±17 | 136±18 | 0.698 |
| Body mass index, | 29.1±5.3 | 75±11 | 77±11 | 0.356 |
| Frailty score | 3.15±1.14 | 3.07±1.06 | 3.22±1.22 | 0.349 |
| Charlson comorbidity score | 5.83±1.67 | 5.73±1.72 | 5.93±1.63 | 0.396 |
| Creatinine, mg/dL | 1.15±0.82 | 1.14±0.78 | 1.16±0.87 | 0.869 |
| eGFR, mL/min/1.73 m<sup>2</sup> | 66±20 | 67±21 | 65±19 | 0.616 |
| Hemoglobin, g/dL | 13.0±1.6 | 13.1±1.7 | 13.0±1.4 | 0.726 |
| NT-proBNP, pg/mL | 1481 (741-3587) | 1850 (779-4051) | 1328 (702-3095) | 0.127 |
| NYHA I/II | 105 (48) | 46 (42) | 59 (53) | 0.103 |
| NYHA III/IV | 115 (52) | 63 (58) | 52 (47) |  |
| 6MWT distance, m | 306±119 | **285±116** | **327±119** | **0.009** |
| 6MWT % predicted, % | 72.1±25.6 | **65.8±24.7** | **78.3±25.1** | **<0.001** |
| 6MWT distance <80% predicted, n (%) | 133 (60) | **82 (75)** | **51 (46)** | **<0.001** |
| Death, n (%) | 39 (18) | 10 (9) | 29 (26) | **<0.001** |
| Betablockers, n (%) | 181 (82) | 91 (83) | 90 (81) | 0.640 |
| ACEI/ARB, n (%) | 161 (73) | 75 (69) | 86 (77) | 0.146 |
| MRA, n (%) | 59 (27) | 24 (22) | 35 (32) | 0.111 |
| Loop diuretics, n (%) | 101 (46) | 50 (46) | 51 (46) | 0.991 |
| Calcium channel blockers, n (%) | 71 (32) | 32 (29) | 38 (34) | 0.437 |
| **Baseline echocardiographic parameters** |  |  |  |  |
| LVEDD, mm | 49.8±6.5 | 50.4±6.3 | 49.2±6.7 | 0.167 |
| LVMI, g/m<sup>2</sup> | 151±38 | 151±35 | 150±41 | 0.765 |
| LAVI, mL/m<sup>2</sup> | 50.3±15.3 | **47.9±12.8** | **52.7±17.1** | **0.018** |
| LVEF, % | 53.3±10.8 | 52.6±11.2 | 54.0±10.4 | 0.326 |
| GLS, % | 14.3±4.5 | 14.4±4.9 | 14.1±4.1 | 0.605 |
| GWI, mmHg% | 2223±830 | 2280±868 | 2167±790 | 0.310 |
| GCW, mmHg% | 2555±870 | 2604±928 | 2507±810 | 0.409 |
| GWW, mmHg% | 188±114 | 178±117 | 198±110 | 0.187 |
| GWE | 90.6±6.8 | 91±6 | 90±7 | 0.201 |
| TAPSE, mm | 21.2±4.6 | 21.5±4.8 | 20.9±4.4 | 0.361 |
| E/A | 1.12±0.58 | **1.23±0.64** | **1.01±0.49** | **0.004** |
| e' septal, cm/s | 4.65±1.41 | 4.68±1.37 | 4.62±1.45 | 0.769 |
| E/e' | 18.2±8.2 | 18.0±8.2 | 18.4±8.5 | 0.726 |
| TR gradient, mmHg | 33.4±11.8 | **35.2±11.5** | **31.6±11.9** | **0.024** |
| AV mean gradient, mmHg | 52.6±15.5 | 54.0±16.1 | 51.2±14.9 | 0.187 |
| AVAI, cm<sup>2</sup>/m<sup>2</sup> | 0.39±0.11 | 0.38±0.11 | 0.42±0.11 | 0.130 |
| LVSVI, mL/m<sup>2</sup> | 42.6±11.6 | 42.8± 1.3 | 42.5±11.4 | 0.860 |
| LFLG aortic valve stenosis, n (%) | 21 (10) | 11 (10) | 10 (9) | 0.784 |
| **Follow-up parameters** |  |  |  |  |
| ΔNYHA class | -1.16±1.06 | **-1.35±1.01** | **-0.95±1.08** | **0.006** |
| Δ6MWT distance, m | 48.2±77.2 | **102±56** | **-12±48** | **<0.001** |
| ΔNT-proBNP, pg/mL | -2246±4229 | -3002±5137 | -1405±2692 | **0.006** |
| ΔLVEF, % | 2.44±6.34 | **3.30±7.03** | **1.50±5.36** | **0.041** |
| ΔGLS, % | 1.33±2.76 | **1.81±2.75** | **0.79±2.68** | **0.007** |
| ΔGWI, mmHg% | -363±579 | -309±619 | -424±527 | 0.150 |
| ΔGCW, mmHg% | -367±686 | -314±779 | -427±562 | 0.237 |
| ΔGWW, mmHg% | -18±137 | -31±136 | -4±137 | 0.154 |
| ΔGWE | 0.52±6.20 | **1.35±6.77** | **-0.41±5.39** | **0.041** |
| ΔTAPSE, mm | -0.34±4.56 | -0.27±4.74 | -0.40±4.38 | 0.836 |
| Δe' septal, cm/s | 0.39±1.22 | 0.44±1.13 | 0.34±1.33 | 0.577 |
| ΔE/e' | -2.96±5.62 | **-4.16±5.95** | **-1.63±4.91** | **0.001** |
| ΔTR gradient, mmHg | -5.75±12.58 | **-8.20±11.52** | **-3.03±13.20** | **0.003** |
| ΔAV mean gradient, mmHg | -42.0±15.6 | -42.7±16.9 | -41.1±14.0 | 0.456 |
| ΔAVAI, cm<sup>2</sup>/m<sup>2</sup> | 0.66±0.28 | 0.64±0.27 | 0.67±0.29 | 0.441 |
| ΔLVSVI, mL/m<sup>2</sup> | 3.36±11.9 | 2.67±12.0 | 4.13±11.9 | 0.380 |

6MWT=six-minute walk test;A=late diastolic mitral flow velocity;ACEI=angiotensin- converting enzyme inhibitors;ARB=angiotensin II receptor blockers;AV=aortic valve;AVAI=aortic valve area index;Δ=increase at follow-up;E=peak early diastolic mitral flow velocity;e'=early diastolic mitral annulus velocity;eGRF=estimated glomerular filtration rate;GLS=global longitudinal strain;GCW=global constructive work;GWE=global work efficiency;GWI=global myocardial work index;GWW=global work waste;LAVI=left atrial volume index;LFLG=low flow, low gradient;LVEDD=left ventricular end-diastolic diameter;LVMI=left ventricular mass index;LVSVI=left ventricular stroke volume index;MRA=mineralocorticoid receptor antagonist;NT-proBNP=N-terminal pro-B type natriuretic peptide;NYHA=New York Heart Association;TAPSE=tricuspid annular plane systolic excursion;TR=tricuspid regurgitant.

### Poor functional response to TAVR

The absence of significant improvement in exercise capacity after TAVR, defined as an increase in 6MWT distance <40 m, was found in 111 patients (50%). These subjects were characterized by older age, longer 6MWT distance, but lower prevalence of 6MWT distance <80% predicted, more prevalent atrial fibrillation, larger left atrial size, and lower E/A ratio and TR gradient. At follow-up, this group presented lesser improvements in 6MWT distance (by definition), NYHA class, NT-pro-BNP, LV ejection fraction, GLS, GWE, E/e’ ratio, TR gradient (**Table 1**).

A cohort of 133 patients exhibited significantly reduced pre-TAVR exercise capacity as indicated by 6MWT distance <falling below 80% of age, sex and body size predicted values. Among these subjects, 39 (29%) demonstrated poor improvement in exercise capacity after TAVR (change in 6MWT distance <40 m). This subset was characterized by older age, higher prevalence of atrial fibrillation, greater need for post-procedural pacemaker implantation, higher frailty, higher GWW, and lower improvement in GLS with intervention than their counterparts presenting post-TAVR improvement in 6MWT distance >40 m (**Supplementary Table 2**).

The independent predictors of poor functional response to TAVR were higher patient age and GWW, coexistence of atrial fibrillation or diabetes, and predicted 6MWT distance above 80%. Nested models demonstrated that the addition of 6MWT distance expressed as a binary variable with a cut-off value of 80% and then GWW to the model including age, diabetes and atrial fibrillation improved the predictive power for this outcome (**Figure 2**). The analogous series of sequential models in the validation cohort confirmed an incremental predictive value of predicted 6MWT distance above 80% and GWW for poor functional response to TAVR (**Supplementary Table 3**).

**Figure 2.**
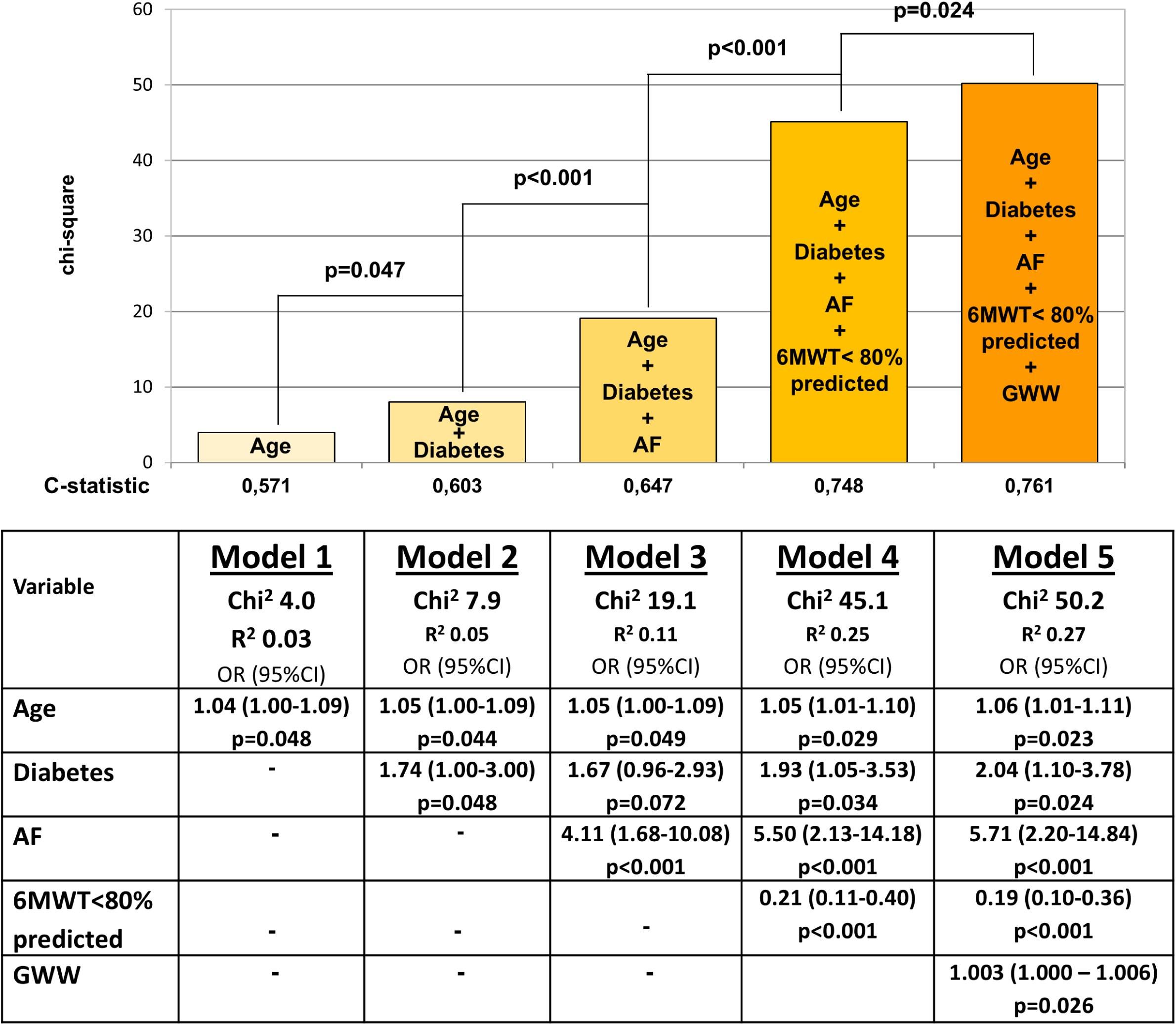
Incremental value of 6MWT and global wasted work for prediction of poor functional response to TAVR.

### All-cause mortality

During a median post-TAVR follow-up of 3.2 years, 39 patients (18%) died. The subgroup with an adverse outcome was characterized by older age, more severe co-morbidity burden, shorter 6MWT distance, and less favorable pre-TAVR echocardiographic profiles, as compared to their peers with event-free survival (**Supplementary Table 4**). A series of multivariable Cox regression models, developed from features showing a univariable association with the outcome, revealed that independent predictors of mortality were older age, reduced GWI or GLS, decreased 6MWT distance, presence of atrial fibrillation, increased patient frailty and increased E/e’ ratio. Due to the high collinearity between GWI and GLS (r=0.89), these two variables could not be included in the same analytical model. However, as assessed by the c-statistic and R-squares, the predictive power of models with GWI was stronger than that of models with GLS (**Table 2**).

**Table 2.** Independent predictors of all-cause death. Series of models with GWI and GLS.

| Models with GWI |  |  |  |  |  |  |  |  |  |  |  |  |
| --- | --- | --- | --- | --- | --- | --- | --- | --- | --- | --- | --- | --- |
|  | Model 1 |  | Model 2 |  | Model 3 |  | Model 4 |  | Model 5 |  | Model 6 |  |
|  | C-statistic=0.794;R<sup>2</sup>=0.631 |  | C-statistic=0.784;R<sup>2</sup>=0.658 |  | C-statistic=0.791;R<sup>2</sup>=0.635 |  | C-statistic=0.784;R<sup>2</sup>=0.628 |  | C-statistic=0.794;R<sup>2</sup>=0.658 |  | C-statistic=0.790;R<sup>2</sup>=0.609 |  |
|  | HR (95% CL) | p | HR (95% CL) | p | HR (95% CL) | p | HR (95% CL) | p | HR (95% CL) | p | HR (95% CL) | p |
| **GWI** | 0.99 (0.99-0.99) | **<0.001** | 0.99 (0.99-0.99) | **<0.001** | 0.99 (0.99-0.99) | **<0.001** | 0.99 (0.99-0.99) | **0.001** | 0.99 (0.99-0.99) | **<0.001** | 0.99 (0.99-0.99) | **<0.001** |
| **6MWT** | 0.99 (0.99-0.99) | **0.008** | 0.99 (0.99-0.99) | **0.005** | 0.99 (0.99-0.99) | **0.017** | 0.99 (0.99-0.99) | **0.001** | 0.99 (0.99-0.99) | **0.011** | 0.99 (0.99-0.99) | **0.014** |
| **Frailty** | 1.34 (1.01-1.77) | **0.045** |  |  | 1.19 (0.88-1.60) | 0.252 |  |  |  |  | 1.32 (0.99-1.74) | 0.051 |
| **AF** | 2.09 (1.05-4.17) | **0.035** | 2.08 (1.04-4.15) | **0.038** |  |  | 2.44 (1.21-4.89) | **0.012** |  |  |  |  |
| **Age** |  |  | 1.07 (1.02-1.13) | **0.009** | 1.06 (1.00-1.13) | **0.036** |  |  | 1.09 (1.03-1.15) | **0.003** |  |  |
| **E/e'** |  |  |  |  |  |  | 1.03 (1.00-1.06) | **0.042** | 1.03 (1.00-1.07) | **0.036** | 1.02 (0.99-1.06) | 0.161 |
| Models with GLS |  |  |  |  |  |  |  |  |  |  |  |  |
|  | Model 1 |  | Model 2 |  | Model 3 |  | Model 4 |  | Model 5 |  | Model 6 |  |
|  | C-statistic=0.776;R<sup>2</sup>=0.558 |  | C-statistic=0.760;R<sup>2</sup>=0.587 |  | C-statistic=0.764;R<sup>2</sup>=0.550 |  | C-statistic=0.764;R<sup>2</sup>=0.559 |  | C-statistic=0.769;R<sup>2</sup>=0.569 |  | C-statistic=0.769;R<sup>2</sup>=0.524 |  |
|  | HR (95% CL) | p | HR (95% CL) | p | HR (95% CL) | p | HR (95% CL) | p | HR (95% CL) | p | HR (95% CL) | p |
| **GLS** | 0.92 (0.86-0.99) | **0.025** | 0.90 (0.83-0.97) | **0.005** | 0.89 (0.83-0.96) | **0.002** | 0.99 (0.99-0.99) | 0.062 | 0.90 (0.84-0.97) | **0.004** | 0.99 (0.99-0.99) | **0.020** |
| **6MWT** | 0.99 (0.99-0.99) | **0.008** | 0.99 (0.99-0.99) | **0.004** | 0.99 (0.99-0.99) | **0.012** | 0.99 (0.99-0.99) | **0.001** | 0.99 (0.99-0.99) | **0.007** | 0.99 (0.99-0.99) | **0.013** |
| **Frailty** | 1.32 (0.99-1.79) | 0.052 |  |  | 1.19 (0.89-1.60) | 0.246 |  |  |  |  | 1.21 (0.89-1.65) | 0.058 |
| **AF** | 2.23 (1.12-4.44) | **0.022** | 2.24 (1.13-4.46) | **0.021** |  |  | 2.59 (1.30-5.19) | **0.007** |  |  |  |  |
| **Age** |  |  | 1.07 (1.01-1.13) | **0.012** | 1.06 (1.00-1.12) | **0.048** |  |  | 1.08 (1.02-1.14) | **0.007** |  |  |
| **E/e'** |  |  |  |  |  |  | 1.03 (1.00-1.06) | **0.041** | 1.03 (0.99-1.06) | 0.065 | 1.02 (0.99-1.06) | 0.163 |

AF=atrial fibrillation. Other abbreviations as in Table 1.

### Cluster analysis

Partition Around Medoids (PAM) algorithm of unsupervised learning with 4 clusters was finally selected as the best approach to patient categorization. The demographic, clinical, and echocardiographic characteristics of these subsets of patients sharing similar features are presented in **Supplementary Table 5**.

- Cluster 1: Fewer comorbidities, lowest NT-proBNP, least impaired 6MWT distance, and best preserved systolic and diastolic function.
- Cluster 2: Intermediate comorbidity burden, mildly elevated NT-proBNP, less impaired LV diastolic function than clusters 3 and 4, and moderate LV systolic dysfunction.
- Cluster 3: Male predominance, severely elevated NT-proBNP, very severe AS often with low-flow low-gradient physiology, profound LV and RV systolic dysfunction, severe LV diastolic abnormalities, and highest pulmonary artery systolic pressure.
- Cluster 4: Oldest age, female predominance, highest comorbidity burden, moderately elevated NT-proBNP, intermediate LV systolic dysfunction, and severe LV diastolic abnormalities.

6MWT distance was more reduced in clusters 3 and 4 than in clusters 1 and 2. Clusters 3 and 4 were significantly associated with increased post-TAVR mortality. However, no significant associations were found between cluster membership and poor functional response to TAVR, either in the overall cohort or in the subgroup with baseline 6MWT <80% predicted (**Table 3**).

**Table 3.** Risk of post-TAVR mortality and poor functional response to TAVR (delta 6MWT <40 m) across the clusters from PAM analysis.

| Cluster | Mortality |  | Delta 6MWT<40m |  | Delta 6MWT<40m in subset with baseline 6MWT<80% predicted |  |
| --- | --- | --- | --- | --- | --- | --- |
|  | HR (95% CI) | p | OR (95% CI) | p | OR (95% CI) | p |
| **1** | - | - | - | - | - | - |
| **2** | 1.93 (0.64-5.86) | 0.25 | 0.98 (0.50-1.91) | >.90 | 1.34 (0.44-4.66) | 0.60 |
| **3** | **5.55 (1.67-18.5)** | **0.006** | 0.74 (0.27-1.98) | 0.60 | 0.64 (0.12-3.04) | 0.60 |
| **4** | **4.54 (1.48-13.9)** | **0.008** | 1.11 (0.49-2.49) | 0.80 | 2.37 (0.76-8.38) | 0.20 |

Compared to Cluster 1

## Discussion

This study demonstrated that in patients with severe aortic stenosis, limited improvement in exercise capacity after TAVR despite the optimal hemodynamic outcome of the intervention is a common finding and is more prevalent with older age, concurrent atrial fibrillation and diabetes, higher GWW, and less impaired 6MWT distance relative to predicted values. The predictive model for poor functional response to TAVR cannot be used to effectively predict post-procedural mortality, showing only limited clinical overlap (age, atrial fibrillation). The 6MWT provides independent and incremental prognostic information for both mortality and lack of significant functional benefit. However, its interpretation differs depending on the prognostic context, with lower absolute values being predictive of mortality, and higher percent predicted values – of lesser improvement in exercise capacity. Myocardial work parameters may enhance prognostic evaluation when planning treatment with TAVR (**Graphical Abstract**).

### Prediction of poor functional response to TAVR

Exercise capacity and tolerance are critical determinants of quality of life. Symptom relief following TAVR is as important an expectation from patients as improved survival. No uniform approach exists to identify patients unlikely to benefit from TAVR. Prior studies identified a number of factors predicting poor symptomatic outcome following TAVR, including age, NT-proBNP levels, presence of atrial fibrillation, chronic obstructive pulmonary disease, chronic kidney disease, patient frailty, and uncorrected mitral valve disease^24–27^. However, no specific patient profile associated with suboptimal post-TAVR functional improvement has been defined.

Our analysis using the predefined definition of poor functional response to TAVR as a change in 6MWT distance <40 m demonstrated that larger post-intervention increases in exercise capacity were associated with shorter pre-TAVR 6MWT distance, which suggests that more profound impairment may provide more space for potential improvement with treatment. Accordingly, greater pre-TAVR limitations in exercise capacity do not preclude meaningful functional recovery following the procedure. Patients with better preserved exercise capacity despite the hemodynamic abnormalities of AS might reflect better temporal adaptation and/or a less disabling disease with less advanced clinical consequences. In this scenario, aortic valve correction may initially result in less functional improvement, but in the long term provide a benefit in preventing functional decline.

In addition, larger gains in 6MWT were accompanied by larger improvements in cardiac function, as evidenced by changes in LV ejection fraction, GLS, GWE, E/e’ and TR gradient. This is consistent with the physiological link between cardiac performance and exercise capacity. When an analysis was confined to patients with significantly impaired exercise capacity at baseline (as defined by 6MWT<80% predicted), GWW and patient frailty emerged as significant associations of poor post-TAVR functional improvement.

A series of multivariable models revealed significant independent contributions to the prediction of poor functional response to TAVR from patient age, presence of atrial fibrillation and diabetes, global wasted work, and degree of 6MWT impairment relative to predicted normative ranges. However, these variables provide an explanation for only 27% of the variation in the outcome, and a search for additional variables would be of value.

### Prediction of mortality

In the present study, we found multiple univariable correlates of mortality. However in the final models, only global work index, 6 MWT distance, patient age, presence of atrial fibrillation, frailty score, and E/e’ showed independent predictive value. The demonstrated associations of these markers with an increased risk of death after TAVR are consistent with the findings from previous studies^8–11, 25, 28–30^. The range of independent prognostic factors reflects the variety of pathophysiological influences on outcome, thus highlighting the multifactorial nature of processes leading to diminished post-TAVR survival.

Moreover, our study demonstrated prognostic superiority of GWI over other metrics of LV systolic function, including GLS. Accordingly, myocardial work assessment may provide more accurate information on myocardial function in the setting of critically increased LV afterload in AS than uncorrected longitudinal deformation. This supports the use of this approach in the diagnostic armamentarium in TAVR candidates.

### Machine learning analysis

Using the PAM method to cluster multidimensional data, we delineated distinct patient phenotypes with varying demographic, clinical and echocardiographic characteristics. These phenogroups differed in terms of post-TAVR mortality. Profiles associated with increased risk of death included males with severe LV and right ventricular abnormalities and pulmonary circulation involvement, low-flow low gradient AS and severely elevated NT-proBNP, as well as older females with moderately increased NT-proBNP and severely impaired LV diastolic filling. Conversely, machine learning-based clustering did not identify specific patient phenotypes associated with the lack of significant gains in exercise capacity after TAVR. These findings support the notion that a poor functional response to TAVR is complex and individually variable, combining both pre- and periprocedural factors.

### Significance of 6MWT

The 6MWT is a simple, reproducible tool reflecting daily activities and functional status. Its advantages include ease of implementation and established prognostic value. Despite this encouraging profile, only a few reports can be found in the literature on the use of 6MWT in pre-TAVR evaluation, which ultimately results in limited data to inform clinical practice^4, 9, 10^. The emergence of 6MWT distance as an independent predictor of post-TAVR mortality and limited symptomatic benefit underscores its potential value in the diagnostic assessment of TAVR candidates.

### Clinical implications

The findings of this study are not intended to inform changes to existing TAVR suitability criteria. However, pre-procedural assessment using the 6MWT and myocardial work indices may support patient profiling by identifying individuals more likely to derive substantial clinical benefit from the intervention. Such information can aid clinical decision-making, particularly in patients with borderline eligibility due to other conditions. Furthermore, clearer expectations regarding post-TAVR improvements in quality of life and survival may help patients make more confident decisions about undergoing treatment.

### Limitations

Several study limitations should be acknowledged. First, the single-center study design might affect the generalizability of the results. Second, given the limited size of our population, we did not develop a predictive score to estimate clinical risk after TAVR. However, our results may provide a basis for further research to address this practical goal. Third, information on the specific cause of death was not consistently obtainable. However, we believe that aggregate data on mortality are of value in this multimorbid, elderly group. Finally, no magnetic resonance data on myocardial characteristics was available, which did not permit inferences about the role of myocardial fibrosis in prognostic outcomes after TAVR.

### Conclusions

This study sheds light on the nature of prognostic associations between post-TAVR outcomes and pre-intervention patient status, indicating distinct features predictive of mortality and failure to improve functional capacity after valve correction. The 6MWT may be useful in predicting the likelihood of both outcomes: lower absolute values are associated with mortality, whereas higher percentage-predicted values are linked to limited symptomatic benefit.

Myocardial work analysis may further strengthen prognostic evaluation. The spectrum of factors predicting poor functional improvement after TAVR does not entirely align with those linked to mortality, suggesting the need for distinct prognostic strategies to assess the risk of these adverse outcomes.

## Data Availability

All data referred to in the manuscript are available on request

